# Beyond Core Duties: A Framework for Reimagining Community Health Worker Contributions to Health Systems

**DOI:** 10.1101/2025.10.21.25336576

**Authors:** Baldeep K. Dhaliwal, Anuradha Nadda, Svea Closser, Melissa Higdon, Madhu Gupta, Shalini Singh, Kerry Scott, Anita Shet

## Abstract

Community Health Workers (CHWs) are essential to global health systems, yet existing research predominately focuses on their core responsibilities, such as maternal and child health, without accounting for the full scope of their labor. This study examines the overlooked dimensions of CHW work through an ethnographic case study of Accredited Social Health Activists (ASHAs) in urban India. Drawing on three months of participant observation and 25 in-depth interviews with ASHAs and health system stakeholders across two sites in Punjab, we identified three interrelated workstreams that define ASHA labor: core duties, secondary responsibilities, and supplementary engagements. Core duties are formally mandated health services implemented through health-related offices. Secondary responsibilities – such as election duties – are often imposed on ASHAs by the health system, but are driven by priorities of non-health ministries. Supplementary engagements involve paid or unpaid work with non-governmental organizations and private actors. Together, these workstreams reveal the extent to which ASHAs are relied upon to fill systemic gaps in under-resourced public systems. These crucial overlapping roles frequently stretch ASHAs beyond their capacity, affecting their ability to deliver on core maternal and child health responsibilities and exposing them to exploitative or informal labor arrangements. Our findings underscore the inadequacy of current frameworks that fail to capture the full complexity of CHW roles. We propose a new conceptual framework to analyze CHW labor holistically, grounded in their personal experiences and the institutional constraints that they face. This framework lays the groundwork for rethinking CHW labor across global contexts, guiding research, policy, and practice aimed at ensuring fair compensation, clearer role delineation, and stronger systemic support for CHWs and for more equitable health systems.

## Introduction

Community Health Worker (CHWs) programs, initially conceived as temporary solutions to health challenges, have evolved into vital components of global health systems (1,2). Predominantly women, CHWs have become essential to the smooth functioning of health systems (3). Literature to date has examined their core duties in health system functioning – their work as community mobilizers for immunization, promoters of maternal and child health, distributors of preventive services, providers of clinical care, and contributors to epidemiological surveillance and record-keeping (4). A systematic review of publications on CHWs from 2005 to 2014 found that the largest proportion of the literature – 35% or 235/678 of included studies – exclusively focused on CHWs’ contributions to maternal, child, and neonatal survival (5). The remaining studies explored CHW contributions to other formal programmatic areas, including vector-borne diseases, non-communicable diseases, tuberculosis, and HIV care.

Despite their essential role in health system functioning and health outcomes, CHWs perform a significant amount of secondary and informal work that has not been explored in the literature. Many studies that explore these types of tasks in any capacity focus on the ‘*task-shifting*’ of specific responsibilities – such as HIV or non-communicable disease management – from overburdened health workers to CHWs (6–10). While studies that explore task-shifting capture one dimension of the additional work placed on CHWs, they fail to account for the broader expansion of CHW roles. As the task-shifting literature limits analyses to specific tasks, it overlooks the full scope of CHWs’ labor and the tensions they navigate in balancing formal mandates, additional tasks, and self-initiated responsibilities. Addressing this gap – as our study aims to do – is crucial for fully understanding CHWs’ contributions and informing policies.

Some literature has explored how frontline health workers manage their core duties alongside the informal roles and secondary responsibilities they take on. In Uganda, drug shop vendors who were constrained by systemic inefficiencies were often found to supplement their formal roles with informal practices to sustain their livelihoods, and meet the needs of community members (11). Similarly, private providers and chemists in India reported navigating formal and informal obligations, filling systemic gaps in ways that blurred regulatory boundaries (12). In Senegal, auxiliary staff and informal brokers reported being assigned tasks outside of their official roles to bridge gaps in under-resourced public systems (13). The CHW literature has also documented systemic tensions when examining how voluntary CHWs balance their assigned health responsibilities with other economic activities to earn an income (14,15). These studies frame these experiences as instances of ‘problem solving corruption’ (16–18), or informal or unauthorized practices that frontline workers engage in; not for personal gain, but to navigate systemic constraints, fill institutional voids, or meet community needs when formal systems fall short. In doing so, the literature overlooks broader dimensions of their roles and the lived realities of healthcare workers’ responsibilities. CHWs frequently assume additional responsibilities beyond their core health-related duties, often undertaking tasks that are assigned informally and lack financial compensation. Given the limited literature on this aspect of CHWs’ work, it is essential to examine how CHWs shape and negotiate their roles – not just as instances of ‘problem-solving corruption’ or strategies for financial survival – but as essential health system contributions.

In this study, we examined the multifaceted roles of CHWs through a case study of Accredited Social Health Activists (ASHAs) – a type of CHW – across two urban sites in Punjab, India. We identified the following three workstreams that aim to capture the full scope of ASHA responsibilities: (1) *Core Duties* – mandated health tasks such as maternal and child health services; (2) *Secondary Responsibilities* – additional duties imposed on ASHAs by the health system, often driven by external agencies; and (3) *Supplementary Engagement* – opportunities with non-governmental organizations (NGOs) or private sector actors that provide financial incentives.

The roles that ASHAs occupy cannot be understood in isolation from the broader systems that shape them. How ASHAs interpret and navigate their responsibilities is deeply influenced by the training they receive, the degree of programmatic support they are afforded, and the implicit and explicit expectations placed upon them by supervisors, community members, and the health system at large. By shedding light on the complexities of these elements of the health system, ASHAs’ roles in balancing formal health responsibilities, ad-hoc assignments, and independent income-generating activities, we aim to offer a more comprehensive understanding and realistic model of CHWs’ roles within health systems.

### Background: The ASHA Program

Established under the National Rural Health Mission in 2005 and later expanded to urban areas through the National Urban Health Mission in 2013, the ASHA program seeks to enhance equitable healthcare access and reinforce health service delivery for underserved communities in rural and urban settings (19,20). The urban component was specifically introduced to improve health outcomes among the urban poor by offering care through a network of Urban Primary Health Centers, Urban Community Health Centers, and urban ASHAs (21). ASHAs play a crucial role in increasing awareness of key social determinants of health, including nutrition, sanitation, and hygiene, while also providing additional targeted support to marginalized groups. Their responsibilities include ensuring continuity of care through home visits, accompanying patients to healthcare facilities, and organizing outreach initiatives such as Health and Nutrition Days. Additionally, ASHAs are expected to collaborate with local organizations to enhance community engagement in health initiatives and empower women as active participants in health advocacy (20). Despite this heavy workload, ASHAs are formally considered to be ‘volunteers’ by the government. While ASHAs receive a small monetary honorarium that differs across states, most of their income comes from performance-based incentives linked to specific maternal and child health services such as antenatal care visits, vaccination visits, or maternal delivery, post-natal follow-up visits, and other tasks.

## Methods

Data were collected over approximately three months across two sites in Punjab, India. The first site was an urban city with approximately 1 million residents, characterized by a growing number of malls, business centers, and newly constructed residential apartment complexes. The second site was a peri-urban setting with a population of approximately 500,000 people, characterized by its unique blend of rural and emerging urban elements, attracting a significant number of migrants for agricultural work and employment in local industrial positions.

This work was conducted by the first author (referred to as “I”) between September-December 2023, with rapport building in late September and initial participant recruitment starting on 04/10/2023 and ending on 07/12/2023. I conducted participant observation with 28 ASHA workers across the two sites, interacting with ASHAs six days a week for several hours each day as they carried out their responsibilities. These activities included door-to-door community outreach, vaccination sessions, hospital visits, survey work, ad-hoc tasks, meetings, and interactions with community members and other health system actors. As an ‘active observer,’ I participated in all ASHA activities, conducted formal and informal interviews, and engaged socially with ASHAs and their families, fostering rapport and gaining a deeper understanding of their experiences. During participant observation I took jottings during interactions, which were later expanded into detailed field notes that captured reflections on daily activities and identified emerging questions for further exploration.

I conducted 25 formal in-depth interviews, supplemented by additional informal interviews. These included 13 interviews with ASHAs and 12 with stakeholders (i.e. policymakers, program implementers, and health officers) across the health system. Interviews ranged from 20 minutes to two hours. Most interviews were conducted in Punjabi, and two stakeholder interviews were conducted in English. All interviews were audio-recorded, except two where stakeholders declined consent for recording; in these cases, detailed field notes were taken instead. I transcribed and translated all interview recordings and reviewed transcripts and field notes weekly with members of the research team to identify data gaps and areas for future probing. After completing data collection, I inductively developed a comprehensive codebook comprising primary codes and sub-codes. This codebook was then applied to all transcripts and field notes using MAXQDA for analysis (22).

Individuals who were observed or interviewed underwent a written informed consent process as outlined in ethical approvals from the Johns Hopkins Bloomberg School of Public Health (IRB #25369), Panjab University (ECR-2308-162), and the Punjab National Health Mission (NMH/PB/CCP/2023/106613-16). In accordance with the *American Anthropological Association’s Statement on Ethics and Principles of Professional Responsibility*, I prioritized ongoing consent as an integral part of the study design and implementation process (23). As ethnography has the potential to blur lines between research and friendship (24), I engaged in ongoing discussions to clarify whether participants intended their shared experiences to be treated as research data or as personal discussions; discussions not considered to be data were excluded from field notes.

I conducted research as a first-generation Punjabi American which significantly impacted the way this research unfolded. I largely benefited from my identity, regularly being told, *“You are one of us, so I’ll help you”.* Further, my identity contributed to my ability to quickly build bonds. Once, after spending 20 minutes with an ASHA, she told me with a shy smile on her face, “*It doesn’t feel like we just met. Does it feel like we just met to you?*” I truthfully told her it felt like we had known each other for a long time. These interactions created a unique ‘*insider-outsider*’ dynamic (25) between me and ASHAs; I was enough of an insider for them to feel comfortable being honest, yet enough of an outsider for them to provide substantial detail to ensure I fully understood what they shared.

## Results

### ** All names changed to maintain participant privacy**

This study leveraged this case study of the ASHAs to identify three distinct workstreams that define the scope of responsibilities for CHWs. The first, *Core Duties*, encompasses the core, mandated tasks integral to a CHWs role, such as maternal and child health services and community outreach. The second, *Secondary Responsibilities*, includes additional responsibilities imposed by the government, but driven by demands from agencies outside of their mandated roles, such as directives from non-health ministries. The third, *Supplementary Engagement*, typically involves private sector actors, NGOs, or other entities offering financial incentives. CHWs often undertake this work to supplement their limited income. Together, these workstreams illustrate the complex ways CHWs engage within the health system (Figure 1). Our research describes these interlocking workstreams in the context of the case study of ASHAs in India.

**Figure 1:**
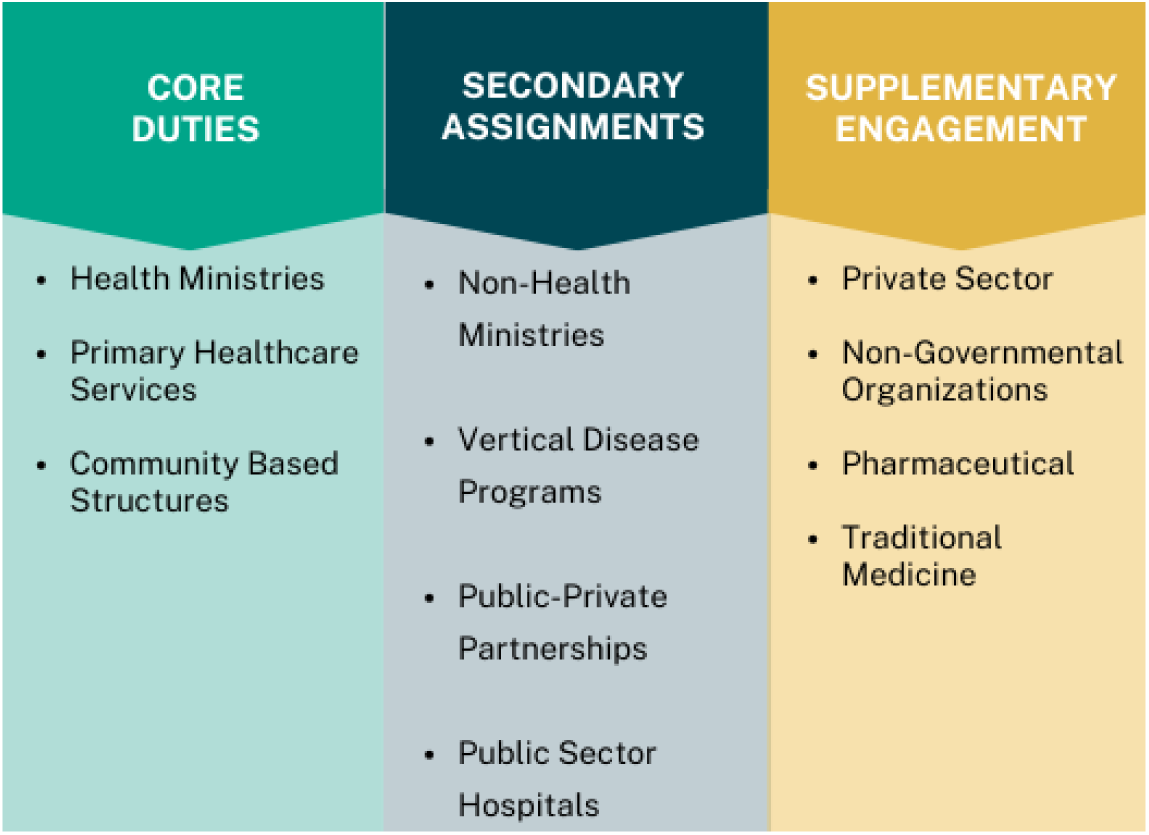
Framework to Assess CHW Engagement Within the Health System.

### Core Duties

As part of ASHAs’ core responsibilities, they were required to engage with the health system in formal, institutionalized ways, acting as conduits for primary healthcare services, maternal and child health services, community-based structures, and other tasks assigned through government health ministries. As a part of these core duties, ASHAs serve as the first point of contact for marginalized populations, ensuring access to essential health services, facilitating health awareness among communities, and providing broad health counseling. Further, their work extends beyond individual health interventions to include community-based engagement with local health committees. ASHAs reported that these core duties were often linked to specific financial incentives, which varied significantly. ASHAs explained that they typically earned nothing for facilitating a community meeting, earned one rupee (less than $0.01 USD) for distributing oral rehydration solution packets, and earned 150 rupees (less than $2 USD) for staying overnight with a mother for a hospital delivery.

Diya, an ASHA who had been working for a year, exemplified the core ASHA duties during one of her routine visits. While walking to conduct household surveys, we walked past a construction site. Diya excitedly explained that this was going to be a mall with brand-name stores and high-end restaurants. She abruptly stopped talking and frowned as she looked ahead at wood and sheet metal structures on the side of the main road.

Although I could see people within the unfinished structures, I thought she was gazing at the remnants of a construction project. I suddenly realized that this was a temporary settlement, something Diya had recognized immediately. As we entered the ‘neighborhood’, there were at least fifty people, most of them young men, women, and small children playing. “*This wasn’t here the last time I came by,*” Diya told me, suggesting that the settlement had been put up quite recently. Recognizing the vulnerability of the families living there, her initial cheerful tone became serious, and she began to systematically approach her work. Pulling out her notebook, Diya wrote down the names of pregnant women and assessed the children’s health and vaccination status.

After observing several children with a similar rash, she gathered their caregivers. “*Chalo! Let’s all go to the health center – Doctor Sahib (sir) will look at them and help,*” she said as she led them to the local health center for treatment. As they walked, Diya explained the services available at the primary health center, “*You won’t have to pay anything for this care – you give your name and your Aadhar (social security and identification) card, and you’ll just wait a few minutes to get your child looked at.*” One mother, visibly relieved, said, *“We didn’t know this was available. No one tells us!”*

For Diya, this outreach was part of her core duties; she was expected to connect underserved communities with existing government health services. She understood these core responsibilities and went about fulfilling them without expectation. Despite their importance and the amount of time they required, these activities did not generate any income for Diya, as they were not linked to a specific incentive.

Participants also reported that as part of ASHAs’ core duties, they were expected to engage with community-based organizations, such as Mahila Arogya Samitis (MAS) or Women’s Health Committees. An ASHA with five years of experience, shared the complexities of this community-based work. *“This is something we’re supposed to do,”* she said, referring to her role in coordinating MAS activities. *“They tell us to explain nutrition, breastfeeding, and sanitation in these meetings, but no one gives us the money we are supposed to have to conduct these meetings. Over time, people have stopped showing up.”* Although ASHAs do not receive direct incentives for implementing MAS meetings, they are supposed to receive funds to facilitate them; sadly, these funds are often not provided. Additionally, there is an expectation that the connections ASHAs build through these meetings will help them earn incentives in the future.

These interpretations of what constitute an ASHAs’ core duties did not occur in a vacuum. Across sites, ASHAs consistently described gaps in both formal training and supervisory support that shaped how they navigated their duties (26). Many ASHAs report working without the mandated eight-day induction, relying instead on peers or overburdened supervisors to guide them. While some learned their responsibilities through informal mentorship, others described feeling underprepared and overwhelmed (26). The absence of structured training and inconsistent support meant that ASHAs adapted their roles with limited formal guidance, often drawing boundaries around their work based on what they were told by their supervisors, or what they felt capable or obligated to do, rather than a clear understanding of programmatic expectations or specific ASHA guidelines.

ASHAs’ deep community ties and their ability to navigate complex health needs made them highly effective in their core responsibilities; in turn, this success in building trust and facilitating access to care led to increased reliance on them for tasks beyond their official mandates.

### Secondary Responsibilities

In addition to their assigned core duties, ASHAs were often tasked with additional responsibilities, and they were promised small incentives as compensation. Government officials outside the Ministry of Health and Family Welfare – where ASHAs are based – frequently delegated extra work to them, including supporting population surveys, election efforts, and educational initiatives. This over-reliance on ASHAs stemmed from both their strong community ties and the essential role they played in service delivery. Several external factors also contributed to this dependence on ASHAs, including chronic workforce shortages across other sectors, the expansion of government health programs without corresponding investments in personnel, and the expectation that ASHAs would bridge gaps in service delivery due to their embeddedness within communities. These secondary tasks were routinely assigned under the assumption that ASHAs’ voluntary status and modest honorariums made them readily available and flexible.

A district-level officer captured this overreliance of ASHAs vividly. As I sat in her office for an interview, she began listing the expansion of ASHAs’ tasks beyond their core responsibilities, counting them off on her fingers, “*On top of maternal and child health work, they have so much more. One, they have to do outbreak investigation – dengue, measles, anything.*” She stopped, frowned, and looked at me. “*You should be writing this down,*” she said. I grabbed my pen and started. Pleased, she continued. *“Next, they have all this survey work for the health center. Then they expect her – a woman – to go talk to men about the dangers of crop burning. They have to also do election work whenever the government says. And even now, they’re the ones being told to go and do the implementation of all these government schemes [programs]*.” Her frustration was palpable: *“ASHAs should be about maternal and child health. When the ASHAs were appointed, they were appointed by the National Health Mission. So why are they being asked to do all this other work?*”

As this district officer shared, non-health Ministries leveraged ASHAs’ connections to the community to complete other tasks. Deepti, a peri-urban ASHA, shared she had recently been tasked with educating people who had small plots of land against crop burning. Even though crop burning was the responsibility of the Ministry of Agriculture, the ASHAs’ supervisor told her it was her “*duty*” to educate people against this, as there were health implications. Resigned to this new responsibility, she explained, “*They told me it’s causing pollution and affecting people’s lungs. So, now it’s my job to tell people to stop burning their plots*.” Another ASHA spoke about work that was assigned to them by the Election Commission of India, saying, *“You know the 2022 election? Our district officer made us go to the polling location at 5:30am. They said because people needed masks and sanitizers this was health-related, so it’s our duty.”*

In addition to supporting non-health ministries with their work, stakeholders reported that ASHAs were regularly asked to support vertical disease programs. A state-based health officer expressed frustration with this, as vertical programs had their own staff, and should not need the ASHA to complete day-to-day support. “*What are all these other people doing?”* she angrily asked in an interview. *“All these other people in vertical programs can do the work, but they just say have the ASHA do it.”* ASHAs were similarly frustrated with their engagement in vertical programs. “*We go and help the patients – we have no choice, and we don’t always get an incentive for this. We just have our orders from above. We can’t even say anything back. They just say, ‘this is the ASHAs work’*.”

ASHAs are often expected to reconcile the demands of core duties and secondary responsibilities on the ground. The success of these processes and outcomes vary due to the limited time, resources and agency granted to the ASHAs. During participant observation with Priyanka, an ASHA who had only been in her role for four months, we walked to a three-level house which had been converted into three apartments. Before ringing the bell to the apartment, Priyanka warned me, “*She’s a bit rude*. *She told me to stop coming so often. I [already] had to go for her antenatal care visit, then they sent me to do a dengue survey, then to fill out a form to see if anyone had ‘sugar’ (diabetes) or ‘pressure’ (hypertension), and now they’re sending me to tell her to register for this insurance program.*” Priyanka’s prediction was correct – the resident sighed as she opened the door, saying, “*I told you last time, you should call. Please don’t just show up*.” Priyanka nodded apologetically, tugged my arm, and we quickly walked away.

Although their strong community ties make them ideal messengers for various government initiatives, the increasing delegation of responsibilities outside their core duties raises concerns.

### Supplementary Engagement

Participants further reported that ASHAs are drawn into supplementary forms of engagement with NGOs, private sector actors, and alternative medicine providers. Some of these engagements are structured and generally accepted, such as when NGOs recruit ASHAs to assist with piloting interventions, conducting community outreach, participating in training sessions, or leading data collection efforts. These activities are often viewed by those in the health system as ‘acceptable tasks’ for ASHAs to take on, since they ultimately serve the community.

When asked about compensation for work with NGOs, one ASHA explained, *“Sometimes they give us a little money or small gifts, like a sari. But other times they promise payment and never give it to us.”* These interactions with NGOs, while presented as opportunities for ASHAs, often fail to deliver tangible benefits; they instead add to ASHAs’ workloads without clear accountability or oversight.

Other supplementary arrangements, particularly those involving private health providers or alternative medicine practitioners, primarily serve to supplement ASHAs’ incomes rather than benefit community health. As these engagements are typically viewed in a negative light by those in and out of the health system, they are often done secretly with no oversight. Local pharmacists and other providers occasionally engaged ASHAs as intermediaries to sell their products. One ASHA recounted how a ‘*medicine man*’ had paid her to refer patients to his stall. *“I would send people to him for cheap medicines, and he gave me part of what he earned [from selling to them],”* she explained. However, the informal nature of this arrangement left her vulnerable; when his stall shut down, he disappeared, owing her unpaid commissions that she was depending on to supplement her income.

ASHAs were also reported to engage with the private sector. A state-level stakeholder described how ASHAs form supplementary partnerships with private health centers to make ends meet, *“The government doesn’t pay them enough, so they end up working with private centers. These centers give them a cut for referring patients*.” Rather than expressing frustration at this arrangement, the stakeholder acknowledged it as an adaptive response to a system that fails to meet ASHAs’ financial needs, and it forces ASHAs to seek out informal arrangements to supplement their income.

These supplementary engagements highlight the precarious position ASHAs navigate as they balance their own financial survival with their professional responsibilities.

### Unintended Consequences on Core Duties: Impact of Overburdening ASHAs

The overlap of core, secondary, and supplementary workstreams significantly impacted ASHAs’ ability to focus on their primary maternal and child health responsibilities. While core responsibilities are central to the role of ASHAs, secondary and supplementary duties often stretched them beyond capacity.

ASHAs reported that their supplementary work with local NGOs required additional time – often with a promise of payment that was not always met. These commitments frequently took time away from their core duties, with one ASHA reporting, “*They make us do trainings for their work, which takes up our time*.” As ASHAs were diverting their attention to this work, they often had less time for essential duties, such as checking on expectant mothers that relied on them for antenatal care and referrals.

A district officer similarly expressed concern that secondary responsibilities placed undue burdens on ASHAs, and it inadvertently was contributing to maternal deaths. While these deaths were infrequent enough to escape an outsider’s notice, as an insider, she recognized their significance because they occurred in areas that hadn’t reported a maternal death in decades. Although the specific causes behind the maternal deaths remained unclear, her personal investigation revealed a common thread – ASHAs consistently cited their expanding workload as a key factor in why they weren’t able to identify vulnerable pregnant women. “*ASHAs say they’re being tasked with crop-burning education, population surveys, TB work, and even election duties, on top of their ASHA responsibilities,*” she said, frustrated. “*As a result, they can’t properly focus on maternal and child health, which is their primary role.”*

The district officer’s concerns felt especially relevant during my participant observation with Rani. For two months, Rani had been going door-to-door in her expansive area to register families for *Ayushman Bharat* – a government-funded health insurance program – a task assigned to her as a secondary responsibility. During this time, two women who also fell in Rani’s assigned area gave birth at home instead of at a hospital; these were the first home births in years in this area. When Rani’s Auxiliary Nurse Midwife (ANM) supervisor questioned her about it, Rani defended herself saying, “*How can I be expected to catch them when the last two months have been focused on the Ayushman registration? They told me they weren’t that far along, and I didn’t have time to check. What was I supposed to do?*” While the ANM worker grew sympathetic, she emphasized that maternal care must remain a priority, despite other tasks. “*The priority must be mothers,”* she gently explained to Rani*. “The more home births we start to have, the more chance that someone can die. We can’t have maternal deaths in this area. All trust will be gone.*”

## Discussion

Our findings underscore the need for a nuanced understanding of the broad contributions that ASHAs make to the Indian health system and a willingness to explore the impact of overburdening on the primary healthcare system. While researchers and practitioners have long anecdotally noted that CHWs take on responsibilities beyond their core duties, these contributions are frequently under-researched or insufficiently examined in the literature. Much of the global literature on CHWs, as well as ASHAs, centers on their core health duties, such as health education, community outreach, and connecting community members to health services (4,5,27). This narrow focus obscures the full range of their labor and the structural tensions they navigate.

Existing frameworks that attempt to broadly define CHW roles – such as the WHO Guidelines on Health Policy and System Support for CHWs – primarily focus on CHWs’ integration into public health systems. Newer frameworks that attempt to broaden the scope of CHW roles, still largely focus on maternal and child health (28). In limiting the focus of CHW responsibilities, this overlooks the informal, adaptive, and cross-sectoral dimensions (29) that constitute additional work CHWs are expected to complete. Our proposed framework extends prior efforts to conceptualize CHW roles, exemplified by Glenton et al (30). Their synthesis in the 2021 supplement *Community Health Workers at the Dawn of a New Era* (31), published by Health Research Policy, provided a foundation for understanding the range of CHW activities across task-based work such as prevention, treatment, and social support. By situating CHWs’ responsibilities within our three interrelated workstreams, we offer a complementary, labor-centered perspective that reveals how CHW roles expand in response to systemic gaps and financial precarity. In doing so, we aim to expand existing frameworks to interrogate the structural reasons behind role expansions.

The existing literature highlights that CHWs are leveraged to fill systemic gaps – at times supporting underfunded programs, addressing staff shortages, or taking on ad-hoc assignments (32–36); when highlighted, these instances are often framed as isolated adaptations or temporary coping strategies (12–15). We argue that these are not isolated adaptations, but systemic features of health systems that have come to rely on CHWs to absorb persistent gaps in service delivery. Our findings call for a more comprehensive understanding of how overextension is built into system design: mapping how responsibilities are distributed, how institutions normalize flexible labor, and how CHWs navigate and negotiate these evolving roles. To address this gap, we designed a framework that used bottom-up strategies to incorporate these dimensions, offering a more accurate lens through which to understand the diverse, adaptive, and often precarious nature of CHW labor. While rooted in the Indian context, it is likely highly relevant across diverse settings, providing a starting point for comprehensive research that recognizes and compensates the full spectrum of CHW responsibilities. This also aligns with broader calls for reform across South Asia, where CHW programs are being reimagined to better integrate with primary health care and respond to emerging health system needs (37). By distinguishing between core duties, secondary assignments, and supplementary engagements, this framework provides an analytical tool for assessing how CHW labor is structured, how CHWs interpret their increasingly complex roles, how responsibilities evolve under different governance systems, and provides an approach to identify and compensate these contributions.

Our findings emphasize that secondary and supplementary assignments are not marginal additions to the core responsibilities that are covered extensively in the literature. Ministries and government agencies increasingly rely on CHWs to implement initiatives, such as pollution awareness and election logistics, as secondary responsibilities. These assignments show how CHWs are viewed as flexible labor, or the default workforce for broad public sector initiatives (38). In doing so, this stretches CHW capacity without adjusting compensation, training, or role clarity. Simultaneously, many CHWs who seek out supplementary engagements with NGOs or private actors can be exposed to exploitative arrangements, irregular payments, and potential conflicts with public health mandates. Left unregulated, both secondary and supplementary engagements compromise CHW alignment with public health priorities and can erode CHWs’ financial security. These added burdens often exacerbate the very vulnerabilities CHWs share with the communities they serve – such as food and housing insecurity – and can contribute to psychological stress, burnout, and professional devaluation (39).

Addressing these tensions demands a structural shift in how CHW labor is recognized at the health system and at the global governance level. As long as CHWs are classified as “volunteers” – while being expected to perform essential services – they may continue to seek supplemental work to survive. Governments that wish to reduce the negative consequences of secondary and supplementary engagements may wish to confront this contradiction directly by addressing CHWs’ standing in the health system. By designating CHWs as volunteers rather than formal employees, the state avoids obligations related to compensation, benefits, and employment protections (40). However, this is a double-edged sword: the informal designation exempts CHWs from rigid institutional mandates, which creates conditions that allow them to pursue supplemental work. Formalizing CHW roles through integration into national health workforce structures – with fair compensation, protections, and standards – offers a path toward resolving this contradiction.

Ultimately, CHWs have become an indispensable, albeit undervalued, pillar of public health delivery. Their labor, spanning core, secondary, and supplementary responsibilities, reflects a broader systemic dependence on flexible, low-cost, and often invisible work. To fully recognize their contributions, researchers must explore and document the full spectrum of CHW engagement, implementers must ground their strategies in CHWs’ lived realities, and policymakers must build robust support systems that reflect the true conditions CHWs navigate. This requires more than top-down policy fixes – it demands embedded approaches that move beyond technocratic framings and capture the social, emotional, and structural dimensions of CHWs’ labor. Without this depth of engagement, we risk simplifying the very systems we aim to improve.

Among the ASHAs we studied, the blurred boundaries between core, secondary, and supplementary roles also signals a shifting health system ecosystem in which ASHAs may be increasingly positioned at the intersection of public and private sector interests. With the rise of national insurance schemes like the Ayushman Bharat– Pradhan Mantri Jan Aarogya Yojana (PMJAY), launched in 2018, many private hospitals can be empaneled into the governments’ PMJAY system, and can then bill the government for care provided to eligible patients. With more private hospitals joining this scheme (41), there is a growing potential for ASHAs to be leveraged by private actors to funnel patients into their care. This raises concerns about the potential redirection of patients towards profit-driven entities, possibly at the expense of equitable care. Policymakers in India would benefit from leveraging this framework to consider the pathways government programs may opens for further privatization of care, fragmentation of health care delivery, and deepening blurred boundaries of ASHAs roles.

While our framework draws on ethnographic fieldwork with ASHAs in India and policymakers may benefit from documenting this full scope of work, the broader patterns we identified are not unique to this setting. Globally, CHWs are similarly tasked with responsibilities that exceed their formal mandates, often without sufficient support or compensation. By distinguishing between core duties, secondary responsibilities, and supplementary engagements, our framework provides a practical tool for global governments, program managers, and donors to map CHW workloads more systematically, identify role inflation, and design interventions that preserve CHWs’ ability to deliver essential services. Without making these distinctions, we risk continuing to collapse state-imposed burdens and informal coping strategies under a generic category of “CHW work,” obscuring the nature of CHW work and the accountability of the system.

Future research must move beyond narrow evaluations of coverage or performance to examine how CHWs navigate overlapping responsibilities, systemic gaps, and competing institutional demands. The framework introduced in this study offers a foundation for such work by clearly delineating the categories of CHW labor that demand further investigation. Comparative and embedded studies across diverse settings are essential to uncover shared patterns, surface context-specific challenges, and inform policy responses.

Reframing how we understand CHW responsibilities is not just about acknowledging their work. It is about understanding the structures that quietly depend on CHWs’ unpaid, underpaid, and invisible labor, and committing to improving the health systems that are supported through their labor.

## Data Availability

All data produced in the present study are available upon reasonable request to the authors

## Acknowledgements

We extend our heartfelt gratitude to the ASHAs, whose resilience, integrity, and unwavering dedication to their communities formed the foundation of this research. Their openness during long interviews, their everyday acts of care, and their willingness to share difficult truths gave us a rare glimpse into a system that often goes unseen. We are honored by their trust and moved by their perseverance. We are also thankful to the Auxiliary Nurse Midwives, Lady Health Visitors, Medical Officers, and district- and state-level health administrators who generously shared their time and insights. We appreciate the support of the Punjab National Health Mission and local health officials for enabling this work.

We would also like to thank the following people who generously read and provided inputs on this article: Drs. Beth Resnick, Henry Perry, Jill Owczarzak, Musarrat Rahman, Nalini Visvanathan, and Peter Winch. We are grateful to Ms. Esmeralda Goncalves who provided an initial design of our framework.

Lastly, we recognize the many hands that made this research possible – the health workers, community members, and researchers whose contributions were vital at every stage. This paper stands as a testament to their shared efforts and to a collective vision for building stronger, more supportive community health systems in India.

